# Disease-specific variant interpretation highlighted the genetic findings in 2325 Japanese patients with retinitis pigmentosa and allied diseases

**DOI:** 10.1101/2023.11.09.23297953

**Authors:** Kensuke Goto, Yoshito Koyanagi, Masato Akiyama, Yusuke Murakami, Masatoshi Fukushima, Kohta Fujiwara, Hanae Iijima, Mitsuyo Yamaguchi, Mikiko Endo, Kazuki Hashimoto, Masataka Ishizu, Toshiaki Hirakata, Kei Mizobuchi, Masakazu Takayama, Junya Ota, Ai Fujita Sajiki, Taro Kominami, Hiroaki Ushida, Kosuke Fujita, Hiroki Kaneko, Shinji Ueno, Takaaki Hayashi, Chikashi Terao, Yoshihiro Hotta, Akira Murakami, Kazuki Kuniyoshi, Shunji Kusaka, Yuko Wada, Toshiaki Abe, Toru Nakazawa, Yasuhiro Ikeda, Yukihide Momozawa, Koh-Hei Sonoda, Koji M. Nishiguchi

**Author notes:** Correspondence: Koji M. Nishiguchi, Department of Ophthalmology, Nagoya University Graduate School of Medicine, 65 Tsurumai-cho, Showa-ku, Nagoya, Aichi, 466-8560, Japan, Koh-Hei Sonoda, Department of Ophthalmology, Graduate School of Medical Sciences, Kyushu University, 3-1-1 Maidashi, Higashi-ku, Fukuoka, 812-8582, Japan. These authors contributed equally to this manuscript (Kensuke Goto, Yoshito Koyanagi).

## Abstract

**Background:** As gene-specific therapy for inherited retinal dystrophy (IRD) advances, unified variant interpretation across institutes is becoming increasingly important. This study aims to update the genetic findings of 86 retinitis pigmentosa (RP)–related genes in a large number of Japanese RP patients by applying the standardized variant interpretation guidelines for Japanese IRD patients (J-IRD-VI guidelines) built upon ACMG/AMP rules and assess the contribution of these genes in RP-allied diseases.

**Methods:** We assessed 2325 probands with RP (n=2155, including n=1204 sequenced previously with the same sequencing panel) and allied diseases (n=170, all newly analyzed), including Usher syndrome, Leber congenital amaurosis, and cone-rod dystrophy (CRD). Target sequencing using a panel of 86 genes was performed. The variants were interpreted according to the J-IRD-VI guidelines.

**Results:** A total of 3564 variants were detected, of which 524 variants were interpreted as pathogenic or likely pathogenic. Among these 524 variants, 280 (53.4%) had been either undetected or interpreted as variants of unknown significance or benign variants in our earlier study of 1204 RP patients. This led to a genetic diagnostic rate in 38.6% of RP patients, with *EYS* accounting for 46.7% of the genetically solved patients, showing a 9% increase in diagnostic rate from our earlier study. The genetic diagnostic rate for CRD patients was 28.2%, with RP-related genes significantly contributing over other allied diseases.

**Conclusion:** A large-scale genetic analysis using the J-IRD-VI guidelines highlighted the unique genetic findings for Japanese IRD patients; these findings serve as a foundation for the clinical application of gene-specific therapies.

## Introduction

Inherited retinal dystrophy (IRD) is a disorder characterized by the degeneration of photoreceptors and the retinal pigment epithelium, leading to symptoms such as night blindness and visual impairment.^1–3^ Retinitis pigmentosa (RP, OMIM 268000) is the most common form of IRD, with a prevalence of approximately 1 in 3000–4000 people; thus, an estimated 2.5 million people worldwide have RP.^1^ ^4^ Although effective treatments for IRD have been limited in the past, recent advancements in gene/variant-specific therapies suggest that IRD might now be amenable to interventions.^5^ Given the progress of genomic medicine, there is a growing interest in the standardized interpretation of variants that can be reproduced by different institutes.

Genetic screening studies for RP have reported variable diagnosis rates, in the range of 37.4% to 66.6% in Western countries^6–9^ and 29.6% to 72.1% in Asia.^10–13^ The rate may be lower in Japan, ranging from 29.6% to 47.9%,^10^ ^14–16^ depending on the report. The variation in these figures could arise from whether the American College of Medical Genetics and Genomics and the Association for Molecular Pathology (ACMG/AMP) guidelines^17^ were applied for variant interpretation or how they were applied, as these guidelines require specification of multiple items that take into account the unique biological and epidemiological characteristics of each inherited disease.^18^ Without predefined guidelines, variant interpretation can differ substantially between institutes, which may contribute to the variability in genetic interpretation of a given variant.^19^ Japan has a different genetic architecture of IRD compared to Western countries and even neighboring countries,^14^ ^20^ ^21^ leading the Japanese Retina and Vitreous Society to publish a modified set of guidelines, the Japanese inherited retinal dystrophy variant interpretation (J-IRD-VI) guidelines. These guidelines are built upon predefined ACMG/AMP rules based on a thorough discussion among the expert members of key genetic laboratories and allow a uniform interpretation of variants tailored specifically for Japanese patients.^22^

In our previous collaborative genetic study using a next generation sequencing (NGS) panel of 86 genes associated with RP, the rate of genetically solved cases (diagnosis rate) in 1204 RP patients was 29.6%.^10^ Unfortunately, the ACMG/AMP guidelines was not applied for variant interpretation and a unique criteria was adopted, which relied heavily on the interpretation of previous reports, without taking into consideration the quality of the evidence provided in the literature. In addition, only typical RP phenotype was included in the study, leaving out the genetic background of allied diseases, which are sometimes difficult to differentiate from RP.

The purpose of this study was to update the genetic findings using the J-IRD-VI guidelines and expand our assessment to include a larger number of patients, especially those with a phenotype that overlapped with RP. This study may serve as a foundation for future gene-variant-specific therapies for IRD in Japan.

## Materials and methods

### Ethic statement and study subjects

This study was conducted in accordance with the principles of the Helsinki Declaration and was approved by the ethical boards of each institute. All participants provided written informed consent at their respective recruiting institute. An overview of the study is shown in figure 1. We collected DNA samples from 2662 patients with IRD from nine Japanese facilities in the Japan Retinitis Pigmentosa Registry Project (2002–2021), namely Nagoya University Hospital (n=790), Kyushu University Hospital (n=706), Tohoku University Hospital (n=336), Yuko Wada Eye Clinic (n=327), Kindai University Hospital (n=203), University of Miyazaki Hospital and Miyata Eye Hospital (n=112), Juntendo University Hospital (n=87), The Jikei University Hospital (n=63), and Hamamatsu University Hospital (n=38). When collecting DNA samples, each sample was assigned an anonymized number by each institute to ensure that the individual subjects cannot be identified by members outside of each institute.

**Figure 1.**
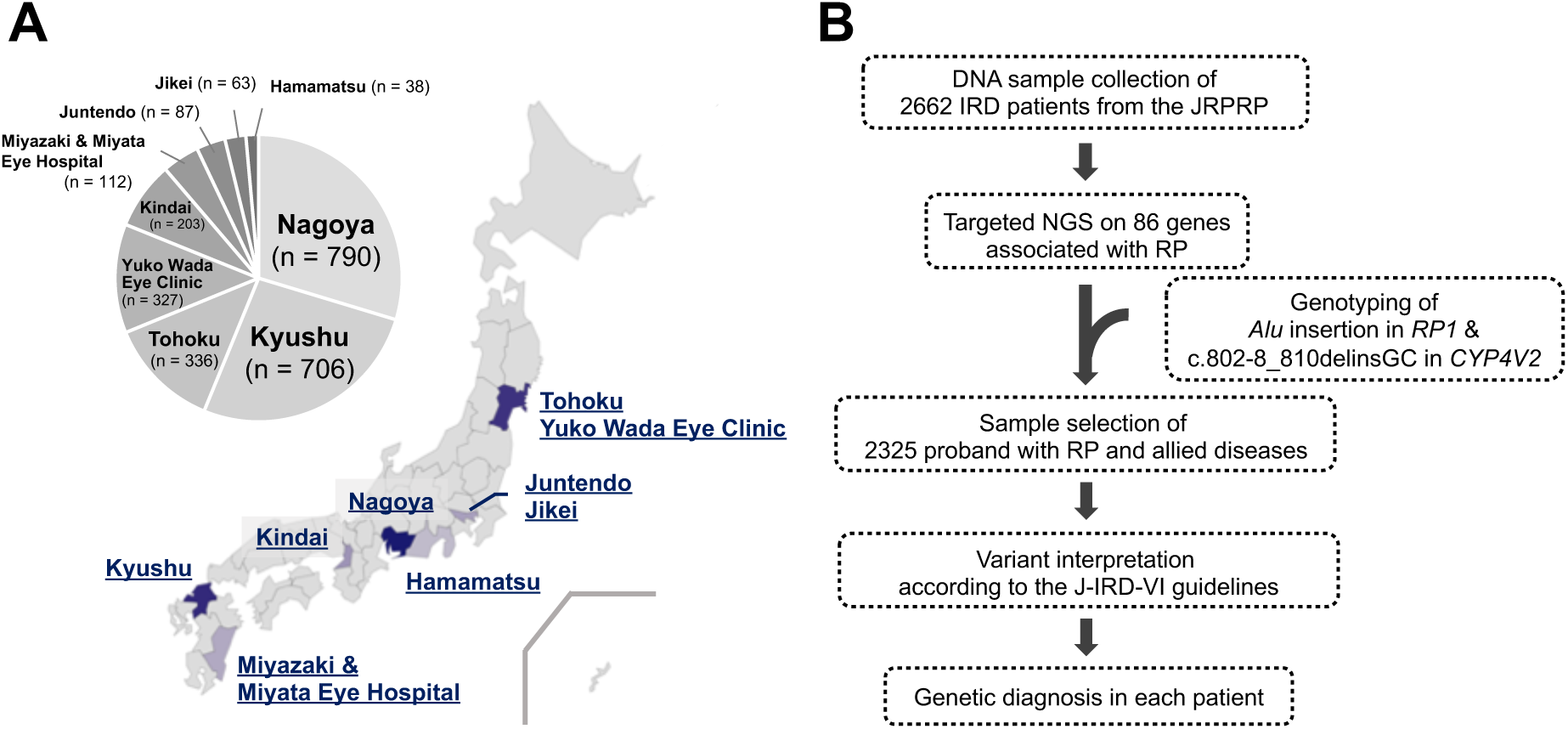
Japanese facilities collecting IRD patients and workflow overview in this study. (A) The proportion of 2662 IRD patients from 9 facilities across Japan and their geographical distribution. (B) The workflow following DNA sample collection. Nagoya, Nagoya University Hospital; Kyushu, Kyushu University Hospital; Tohoku, Tohoku

### Patient selection

From a total of 2662 patients, we selected for analysis patients with RP and patients with allied diseases with clinical findings that overlap with RP, including Usher syndrome, Leber congenital amaurosis (LCA), cone-rod dystrophy (CRD), choroideremia, and Bietti crystalline dystrophy (BCD). Clinical diagnoses were made by well-trained ophthalmologists based on symptoms such as visual acuity loss, visual field loss, photophobia and night blindness, fundus examination, electroretinogram, fundus autofluorescence images, and optical coherence tomography images. We excluded patients (n=258) with IRD with substantially different phenotypes from RP, including macular dystrophy, Best disease, and Stargardt disease. We also excluded related patients (n=80) so that a single patient represented each family. As a result, we enrolled 2325 probands with non-syndromic RP (n=2155, including n=1204 we had previously sequenced with the same sequencing panel^10^) or allied diseases (n=170), such as Usher syndrome (n=32), LCA (n=15), CRD (n=78), choroideremia (n=11), or BCD (n=34).

### Target sequencing for 86 genes

We used a previously established panel of 86 genes associated with RP and allied diseases.^10^ These genes include 83 genes associated with RP that were registered in the Retinal Information Network (RetNet) as of September 19, 2017, as well as three additional genes (*CHM*, *CEP290*, and *IMPG1*) that are major causative genes for other IRDs. Our study was designed to capture the coding regions of the transcripts as well as 2 bp into each exon-adjacent intron and also include a deep intronic variant, c.2991+1655 A>G, in CEP290. Multiplex polymerase chain reaction (PCR)–based target sequencing was performed as described previously.^23^ Briefly, multiplex PCR was performed by designing primers covering all coding regions of the 86 genes. Next, a second PCR test was performed to add dual barcodes to the first PCR product. Finally, the second PCR product was sequenced using a HiSeq 2500 instrument (Illumina, San Diego, CA) to obtain 2 × 151 bp paired end reads with dual 8 bp barcode sequences.

### Analysis of sequencing data

Analysis of sequence data was performed as previous reported.^10^ Sequencing reads were demultiplexed using bcl2fastq2 V.2.20 (Illumina), followed by alignment to the human reference genome (hg19) using Burrows-Wheeler Aligner (V.0.7.17). Post-alignment processing, including indel realignment, was performed with the Genome Analysis Toolkit (GATK) (V.3.7), leading to variant calling via HaplotypeCaller and UnifiedGenotyper, and subsequent integration of variants. Alternative allele frequencies (AF) were calculated using SAMtools (V.1.6) and genotype variants were selected based on frequency histograms and quality filtering. The final set of identified variants was annotated using ANNOVAR (V.3.4) and SnpEff (V.4.3), with the transcripts registered in CCDS Release 24 and refGene. We defined the covered region as a region with bases having ≥20 sequencing reads in the target region. The coverage per base was calculated as the number of covered samples divided by the number of all samples. The cover rate per gene (%) was defined as the average coverage per base of the target region. We successfully sequenced ≥ 98.0% of the targeted regions of the 86 genes in all 2325 samples and used all samples for further analysis. The average read depth (±SD) was 1410 (±359) per sample. We called variants in the regions with ≥20 read depth, which included 99.65% of the targeted regions (online supplementary table S1). We confirmed the presence of pathogenic variants through visual inspection using the integrative genomics viewer (IGV), making manual corrections for annotations when multiple alternative alleles or single-nucleotide variants with deletions were identified at the same site. This led to the reassignment of annotations for 10 variants across four genes (see online supplementary figure S1 and table S2).

### Genotyping of the founder variants in *RP1* and *CYP4V2*

In addition to NGS panel of 86 genes, we analyzed each of two variants associated with IRDs^24–26^ in a different way, with *Alu* insertion in *RP1* and c.802-8_810del17insGC in *CYP4V2*; these genes are not detectable with our NGS panel. To screen *Alu* insertion in exon 4 in *RP1* in all patients, we searched for *Alu* sequences in FASTQ files obtained by NGS using a grep search program, as previously reported.^27^ We selected patients for whom we had either detected *Alu* sequences in at least one read or who were heterozygous for pathogenic variants in *RP1* as the subjects for genotyping. Genotyping was performed using an optimized PCR-based method for the selected patients as previously described.^24^ Among the subjects, 23 patients whose genotyping of *Alu* insertion was previously reported by us^24^ were referenced for those results. To screen c.802-8_810del17insGC in *CYP4V2* in all patients, we developed a grep search program for FASTQ format files. We searched for a unique 20-base sequence containing c.802-8_810del17insGC in *CYP4V2* using the grep command in Linux (online supplementary table S3), and the AF is calculated as (alternative allele read count) / {(alternative allele read count) + (reference allele read count)} in all patients. Allele frequencies ranging from 0.25 to 0.75 were defined as heterozygotes, while frequencies ranging from 0.85 to 1.0 were defined as homozygotes.

### Variant interpretation

Among the 86 genes in our NGS panel, two genes that had previously been reported as associated with RP but whose pathogenicity has been questioned by recent studies^28–30^ were excluded from our analysis, resulting in a final set of 84 genes for interpretation of pathogenicity. We interpreted the variants according to the J-IRD-VI guidelines^22^ published by the Japanese Retina and Vitreous Society. A detailed description of the evaluation procedures is provided below. The automated application AutoPVS1 (http://autopvs1.genetics.bgi.com/)^31^ (accessed on 16 August 2022) was used to evaluate the PVS1 criteria for null variants. As defined in the ACMG guideline criteria for hearing loss^31^, we defined exons containing null variants with an AF exceeding 0.3% in the Genome Aggregation Database (gnomAD)^32^ as lacking functional significance. For 23 null variants that output errors (as not “null variants”) due to differences in the definition of the coding region in AutoPVS1, the PVS1 criteria were evaluated manually. To assess the frequency of variants in general populations (BA1, BS1, PM2), we used the gnomAD total population, the gnomAD population max, the Human Genetic Variation Database (HGVD), and Tohoku Medical Megabank Organization (ToMMo) 8.3KJPN. If the highest AF in any of the four databases met the cut-off value, the respective criteria were applied to variants. BA1 was not applied based on an expert opinion for variants previously reported as being pathogenic by multiple groups but meeting the BA1 cut-off value (online supplementary table S4).^33–38^ To calculate odds ratios for minor alleles in the PS4 criteria, ToMMo 38KJPN for autosome and 14KJPN for chromosome X were used as ancestry-matched controls. To evaluate PM3, we counted the number of probands with heterozygous pathogenic variants and those with homozygotes from both our subject patients and the prior literature listed in the Human Genome Mutation Database Professional (HGMD Professional, 2022.2). The strength of PM3 was assessed by summing the counts of these two sets of probands. In the computational evidence category (PP3, BP4, BP7), we employed predictive tools: REVEL^39^ for predicting the pathogenicity of missense variants and Splicing AI^40^ for predicting splice site changes. The PP5 criteria were evaluated based on ClinVar (accessed on 20 August 2022). The BP2 criteria were applied to variants founded in cases with homozygous pathogenic variants within the same gene.

### Criteria for genetic diagnosis

We performed a genetic diagnosis using the variants regarded as “pathogenic” or “likely pathogenic” by the J-IRD-VI guidelines, as well as the two founder variants, *Alu* insertion in *RP1* and c.802-8_810del17insGC in *CYP4V2*. We applied the genetic inheritance forms defined in RetNet for each gene, following the classical Mendelian inheritance pattern and attempted to determine the causative variant and make a genetic diagnosis in each case as previously reported.^10^ If a gene was registered on RetNet in multiple inheritance forms, we determined the genetic inheritance form of each variant based on the prior literature. Furthermore, when a patient had pathogenic variants in multiple genes leading to a genetic diagnosis, all the genes were listed as potential causative genes, rather than selecting just one. Finally, we regarded patients whose causative genes were determined as “solved” and patients whose causative genes could not be determined as “unsolved”.

## Results

### Results of variants interpretations of 86 genes in 2325 patients

The clinical characteristics of the patients are summarized in table 1. In the RP patients, the modes of inheritance as obtained from clinical records were as follows: AD in 284 (13.2%), AR in 410 (19.0%), XL in 34 (1.6%) and sporadic in 1182 (54.8%) (table 1). A comprehensive list of variant interpretation using J-IRD-VI guidelines is shown in online supplementary table S5. We identified a total of 3564 variants, including 2143 missense variants, 169 nonsense variants, 168 frameshift indels, 40 non-frameshift indels, 53 canonical splice site variants, and 973 synonymous variants.

**Table 1.** Characterisics of 2325 Patients with RP and allied diseases.

| Diagnosis | RP | Usher syndrome | LCA | CRD | Choroideremia | BCD |
| --- | --- | --- | --- | --- | --- | --- |
| Patients, n | 2155 | 32 | 15 | 78 | 11 | 34 |
| Sex (Male/Female) | 1020/1135 | 12/20 | 6/9 | 46/32 | 11/0 | 9/25 |
| Age, years | 54.2 (4-93) | 45.5 (10-80) | 25.3 (3-57) | 55.4 (3-84) | 48.5 (3-84) | 55.8 (27-82) |
| Visual acuity (logMAR), n (%) |  |  |  |  |  |  |
| ≤ 0.5 | 1302 (60.4) | 19 (59.4) | 2 (13.3) | 20 (25.6) | 9 (81.8) | 20 (58.8) |
| 0.5 <, ≤ 1.3 | 381 (17.7) | 2 (6.3) | 4 (26.7) | 38 (48.7) | 1 (9.1) | 7 (20.6) |
| > 1.3 | 391 (18.1) | 9 (28.1) | 8 (53.3) | 18 (23.1) | 1 (9.1) | 6 (17.6) |
| Unknown | 81 (3.8) | 2 (6.3) | 1 (6.7) | 2 (2.6) | 0 (0) | 1 (2.9) |
| Inheritance form, n (%) |  |  |  |  |  |  |
| AD | 284 (13.2) | 0 (0) | 0 (0) | 9 (11.5) | 0 (0) | 2 (5.9) |
| AR | 410 (19.0) | 12 (37.5) | 6 (40) | 8 (10.3) | 0 (0) | 12 (35.3) |
| X linked | 34 (1.6) | 0 (0) | 0 (0) | 0 (0) | 6 (54.5) | 0 (0) |
| Sporadic | 1182 (54.8) | 16 (50) | 9 (60) | 59 (75.6) | 3 (27.3) | 19 (55.9) |
| Unknown | 245 (11.4) | 4 (12.5) | 0 (0) | 2 (2.6) | 2 (18.2) | 1 (2.9) |
| Consanguinity of patients, n (%) | 152 (7.1) | 2 (6.3) | 8 (53.3) | 4 (5.1) | 0 (0) | 3 (8.8) |
| Visual acuity is derived from the right eye. The values of the decimal visual acuity were converted into the logarithm of the minimum angle of resolution (logMAR) units. The inheritance forms were determined based on clinical examination and interview. RP, retinitis pigmentosa; LCA, Leber congenital amaurosis; CRD, cone-rod dystrophy; BCD, Bietti crystalline dystrophy. AD, autosomal dominant; AR, autosomal recessive. |  |  |  |  |  |  |

The detected variants were interpreted according to the J-IRD-VI guidelines using an in-house program. As a result, 211 variants were interpreted as pathogenic, 313 variants as likely pathogenic, 1880 variants as variants of unknown significance (VUS), 516 variants as likely benign, and 644 variants as benign. Among the 524 variants interpreted as pathogenic or likely pathogenic, 209 (39.9%), 249 (47.5%), and 290 (55.3%) had not previously been reported in our earlier study of 1204 patients with typical RP, HGMD, or ClinVar, yielding 107 new disease-associated variants. This included 71 variants considered VUS or benign variants in the previous study. At the same time, 118 variants considered pathogenic or likely pathogenic were reassigned to the VUS or benign categories. Thus, only 245 out of 363 pathogenic or likely pathogenic variants (67.5%) in the previous study maintained the same interpretation in the current study.

### Genotyping results of *Alu* insertion in *RP1* and c.802-8_810del17insGC in *CYP4V2*

Two frequent variants in *RP1* and *CYP4V2* are known to be associated with RP and allied diseases and are missed by the standard analysis pipeline; thus, we analyzed them in a different way. To look for the known common 328bp *Alu* insertion [c.4052_4053ins328, p.(Tyr1352Alafs)] in *RP1,*^24^ ^26^ we conducted screening of FASTQ files using a grep search program^27^ in all patients. *Alu* sequences were detected in at least one read for 24 patients. Then, we selected a total of 138 patients for whom we either detected *Alu* sequences in at least one read or who were heterozygous for pathogenic variants in *RP1* as the subjects for genotyping. These samples were further analyzed with a PCR-based method. We found that 24 patients were heterozygous for the *Alu* insertion and 10 were homozygous. Among the 24 patients who were heterozygous carriers of *Alu* insertions, 18 were found to have another pathogenic variant in *RP1*. As for the splice site variant c.802-8_810del17insGC in *CYP4V2*, which is associated with BCD^25^, we screened all patients by the searching for a 20-base sequence encompassing the variant region in FASTQ files using software. This revealed that 22 patients, including 16 with RP and 5 with BCD, were heterozygous and 16, including 13 with BCD, were homozygous for this change (online supplementary table S6). Among the 22 patients who were heterozygous, 5 with BCD and 1 with RP were found to have another pathogenic variant in *CYP4V2*.

### High-frequency pathogenic variants in solved and unsolved patients with RP

Figure 2 summarizes the proportion of causative variants detected in the solved patients with RP (n=831) and pathogenic or likely pathogenic variants detected in unsolved patients with RP (n=1324) (figure 2B). In particular, [p.(Ser1653fs), p.(Gly843Glu), and p.(Tyr2935*) in *EYS*] and *Alu* insertion [p.(Tyr1352Alafs)] in *RP1* were found in 27.2% (226/831), 20.1% (167/831), 11.9% (99/831), and 2.9% (24/831) of solved patients, accounting for 18.2%, 13.2%, 8.1%, and 2.3% of the total alleles of causative variants, respectively (figure 2A). Among unsolved patients, 37.0% (490/1324) were found to carry pathogenic or likely pathogenic variants. Specifically, [p.(Gly843Glu), p.(Ser1653fs), and p.(Tyr2935*) in *EYS*] were detected in 7.3% (96/1324), 5.0% (66/1324), and 3.0% (40/1324) of unsolved patients, accounting for 16.4%, 11.2%, and 6.8% of the total alleles of pathogenic or likely pathogenic variants carried in unsolved patients, respectively (figure 2B).

**Figure 2.**
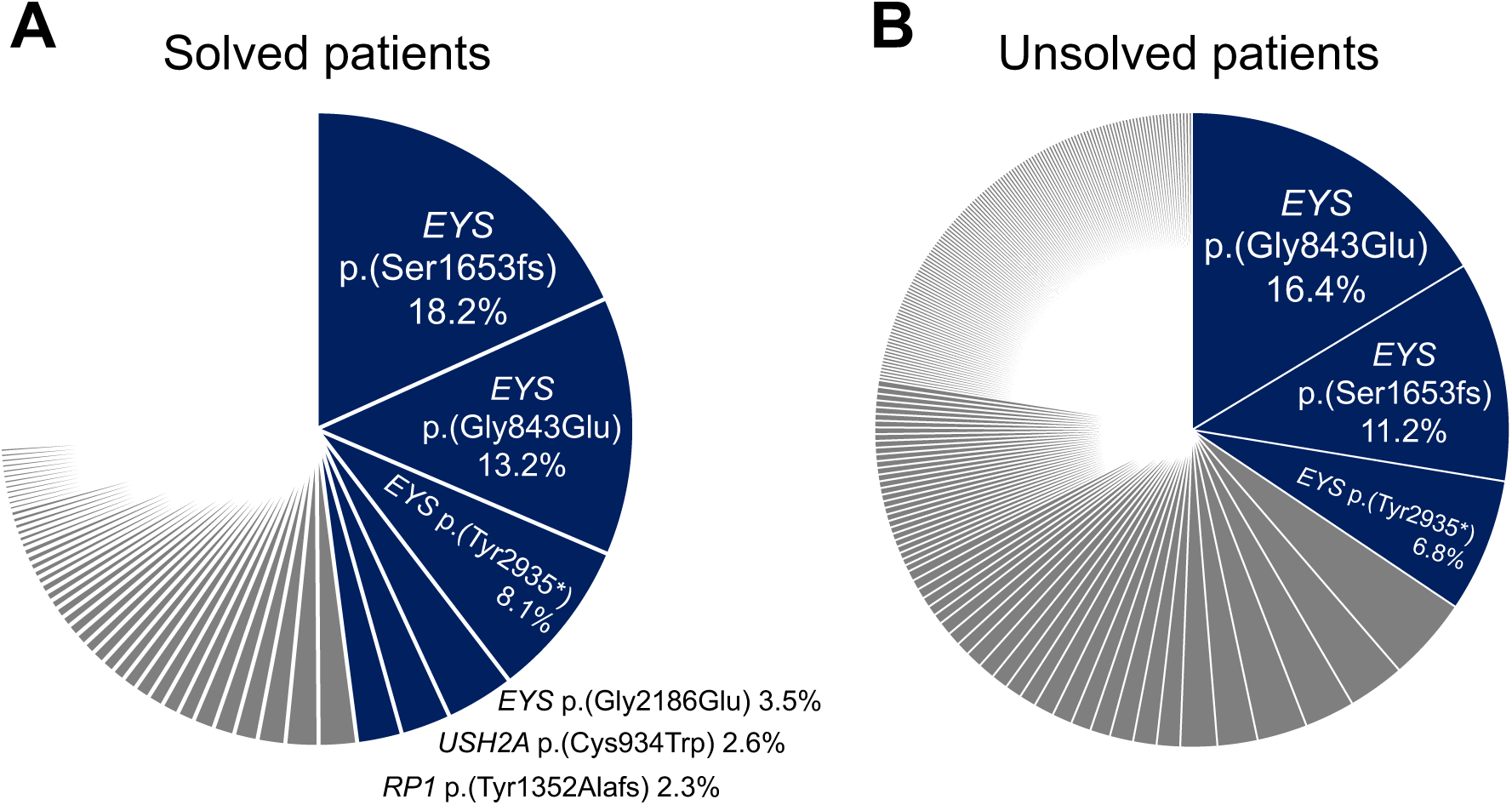
High frequency pathogenic variants in solved and unsolved patients with RP. (A) The proportion of causative variants detected in solved patients with RP. A total of 1485 alleles of causative variants were detected in solved patients with RP. High-frequency variants detected with more than 30 alleles are highlighted in the blue area. (B) The proportion of pathogenic or likely pathogenic variants carried in unsolved patients with RP. A total of 587 alleles of pathogenic or likely pathogenic variants were detected in unsolved patients with RP. High-frequency variants detected with more than 30 alleles are highlighted in the blue area. RP, retinitis pigmentosa.

### Genetic diagnosis with the detected pathogenic variants

The overall genetic diagnosis rate was 38.4% (892/2325) for RP and allied diseases (figure 3). The figure was 38.6% (831/2155) for RP, 25.0% (8/32) for Usher syndrome, 6.7% (1/15) for LCA, 28.2% (22/78) for CRD, 63.6% (7/11) for choroideremia, and 67.6% (23/34) for BCD. The results for causative genes and variants for a total of 2325 patients are summarized in online supplementary table S7. Of note, we detected 15 patients with pathogenic or likely pathogenic variants in two genes, including 14 patients with RP and one patient with choroideremia (online supplementary table S8).

**Figure 3.**
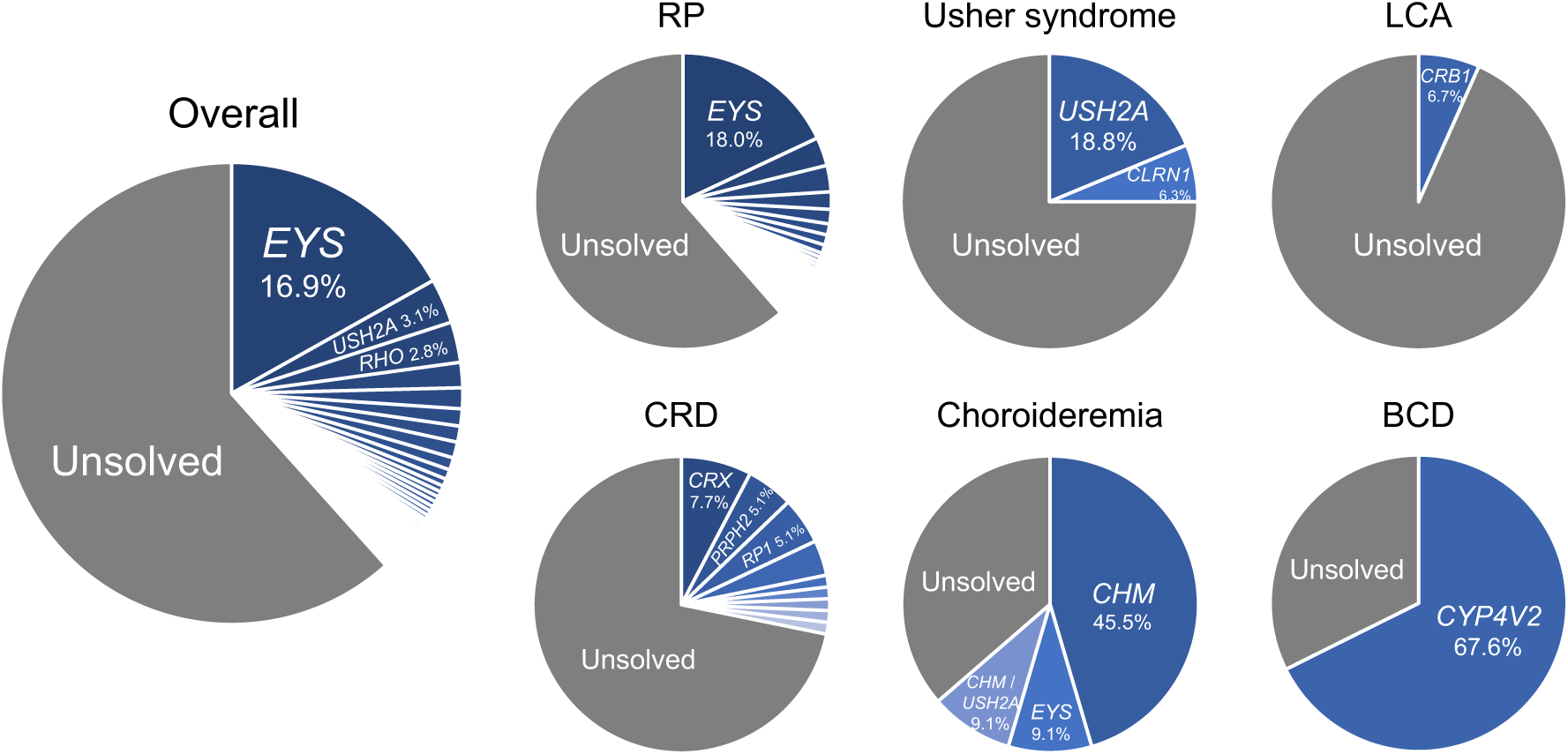
Percentage of solved patients and proportion of causative genes. The pie chart displays the overall and disease-specific diagnostic rates along with the proportion of disease-causing genes. The blue area represents the proportion of genetically solved patients whereas the grey area represents that of unsolved patients. RP, retinitis pigmentosa; LCA, Leber congenital amaurosis; CRD, cone-rod dystrophy; BCD, Bietti crystalline dystrophy.

Among the typical RP patients, 831 were genetically diagnosed with 320 pathogenic or likely pathogenic variants in 49 genes (figure 4A). Pathogenic variants in *EYS*, which has previously been reported to be the major gene responsible for RP in the Japanese population, explained RP in nearly half (388/831, 46.7%) of solved patients, followed by *USH2A* (67/831, 8.1%), *RHO* with dominant inheritance (41/831, 4.9%), *RP1* with recessive inheritance (36/831, 4.3%), and *RPGR* (34/831, 4.1%). The breakdown of solved cases by inheritance pattern based on clinical information was 47.2%, 43.9%, 33.6%, and 41.2% for AD, AR, sporadic, and XL, respectively. When separately analyzed by inheritance pattern of each gene reported in RetNet, *EYS* (388/605, 64.1%) and *USH2A* (67/605, 11.1%) accounted for 75.2% of patients diagnosed with ARRP (figure 4B).

**Figure 4.**
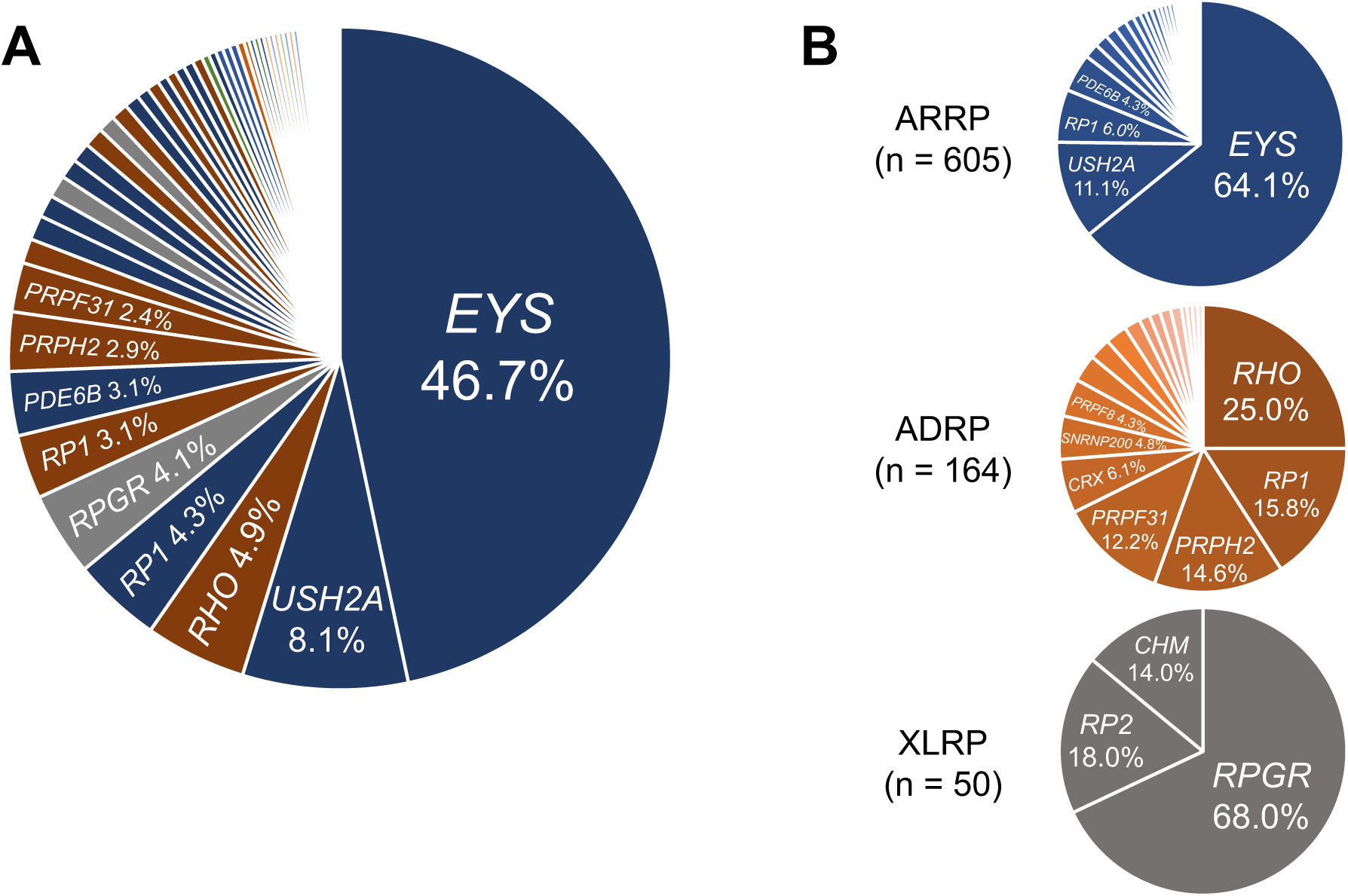
Proportion of causative genes in solved patients with RP. (A) The proportion of causative genes in solved patients with RP. The blue area represents the proportion of AR genes, the brown area represents AD genes, and the grey area represents X-linked genes. The green area represents multiple causative genes. (B) The proportion of causative genes based on the inheritance pattern. The pie chart excludes 12 patients in whom multiple causative genes with different inheritance patterns were detected. AD, autosomal dominant; AR, autosomal recessive; n, number of patients; RP, retinitis pigmentosa.

Meanwhile, three genes (*RHO*, *RP1*, and *PRPH2*) explained more than half of solved cases with ADRP (91/164, 55.5%). In X-linked RP (XLRP), pathogenic variants in *RPGR* were found in the large majority of solved patients (n=34/50, 68.0%).

In Usher syndrome, a total of eight of 32 patients (25.0%) were genetically diagnosed. Six patients had pathogenic variants in *USH2A* and two patients had pathogenic variants in *CLRN1*. In LCA, only one of 15 patients was genetically diagnosed, while no pathogenic variant was detected in the remaining 14 (93.3%) patients, consistent with a previous report indicating little overlap between genes linked to RP and LCA in Japan.^15^ ^41^ In CRD, variants in *CRX*, *PRPH2*, and *RP1* were the most commonly linked to the disease, each accounting for 7.7%, 5.1%, and 5.1% of the disease, respectively. Of the 11 patients diagnosed with choroideremia, pathogenic variants in *CHM* were detected in six patients (54.5%). Variants in the *CYP4V2* gene explained 67.6% (23/34) of patients with BCD, and c.802-8_810del17insGC was detected in 5 compound heterozygotes and 14 homozygotes.

## Discussion

In this study, we performed target sequencing in 2155 patients with RP and an additional 170 patients with allied diseases. This is nearly double the number of RP patients as in our previous study, although 1204 patients from the previous study were also included in the present one.^10^ We interpreted the detected variants according to the J-IRD-VI guidelines which is based upon specified ACMG/AMP rules and tailored explicitly for IRD in the Japanese population.^22^ Our results showed that the genetic diagnosis rate was 38.6% in the RP patients, which is higher than the corresponding rate of 29.6% in our previous work,^10^ which adopted a different variant interpretation pipeline. Our findings are supported by the diagnostic contribution of 107 newly assigned variants not reported in ClinVar, HGMD, or our previous work, which comprised 20.4% of all pathogenic or likely pathogenic variants detected in this study. In addition, extra genotyping of common *Alu* insertion in *RP1* and a splice site variant in *CYP4V2*, both missed by our automated NGS analysis, contributed to the increased diagnosis rate.

Although 1204 patients examined in our previous work were also included in this study, only 67.5% of pathogenic or likely pathogenic variants had the same interpretation in the earlier work and this one. This could largely be attributed to the difference in interpretation criteria; the previous study adopted variant interpretation posted in ClinVar and HGMD. As a result, the rate of contribution of the most prevalent disease genes changed considerably. In particular, the proportion of *EYS* jumped from 30.9% to 46.7% in the current study, which may be explained in part by the recent establishment of a very frequent variant [p.(Gly843Glu) in *EYS*] as pathogenic.^36^ Additionally, through a stringent application of J-IRD-VI guidelines, nearly all variants in *RP1L1*, which was previously the fifth most prevalent RP-related gene in Japan and accounted for 4.8% of all solved RP cases, were excluded in this study due to the high AF in the general population as defined in the J-IRD-VI guidelines. Consequently, the current study has greatly enhanced the understanding of the genetic architecture of Japanese RP patients.

This study also found that 37.0% of the unsolved RP patients carried pathogenic or likely pathogenic recessive variants in one of the 86 RP-related genes. The AF of many of these variants was higher than would be expected in the general population. For example, the AF of p.(Ser1653fs) was 2.49% in unsolved RP patients and is 0.43% in the general population. This may indicate that some of these heterozygous variants play a role in the disease in unsolved cases and that there is a yet-unidentified pathogenic recessive variant *in trans* that escaped the NGS panel sequence and standard analysis pipeline. Indeed, in an earlier analysis, we carried out a genome-wide association study of RP, which showed a peak in the *EYS* locus that surpassed genome-wide significance (P =3.79×10^-10^) and for which no corresponding exonic variant in *EYS* was linked,^36^ indicating that there may be at least one more undiscovered high-frequency variant in this gene. Furthermore, through long read sequencing of 15 heterozygous carriers of an *EYS* pathogenic variant, we identified a large structural variant encompassing multiple exons in *EYS* in 2 cases (13.3%),^42^ suggesting the presence of undiscovered structural variants. Nevertheless, a more complex non-Mendelian inheritance factor may also complicate genetic diagnosis,^43–45^ although the degree of its importance in the context of overall genetic diagnosis remains largely unknown.

This study reports genetic analysis results of IRD cases with phenotypic overlap with RP in a somewhat limited number of patients. We found a significant contribution of RP-related genes in the genetic diagnosis rate of CRD (22 of 78 patients, 28.2%), although the figure was smaller than for RP (38.6%). Unlike RP, variants in *CRX* were the most frequent causes of CRD and *EYS* variants accounted for the disease in only 3.8% of cases. This was in contrast to patients with BCD or choroideremia, both rarely diagnosed as RP, in which variants in *CYP4V2* and *CHM*, respectively, explained most patients. Meanwhile, we reached genetic diagnosis in only one of 15 patients with LCA, consistent with previous reports indicating that genes causing RP and LCA are largely different in the Japanese population.^15^ ^41^ Nevertheless, these findings are important in the era of genomic medicine because many emerging treatments for IRD are developed for each disease gene with less emphasis on the clinical diagnosis itself; thus, understanding the phenotypic spectrum of each disease gene is critical for the clinical application of these treatments.

There several limitations to this study. First, the J-IRD-VI guidelines used for variant interpretation are customized for and validated in only Japanese IRD patients. For example, mainly Japanese databases were designed to assess the rarity or enrichment of a variant in question. This means that the same analysis pipeline may not be applicable to countries outside Japan, even neighboring Asian countries. Nevertheless, since Japan consists of islands geographically isolated from elsewhere, it is unavoidable that there will be unique criteria for a sensitive and specific genetic diagnosis. Second, the RP-related genes analyzed in the current study were selected based on those reported as causing non-syndromic RP in RetNet as of 2017. They are not updated, nor do they comprehensively cover genes reported for phenotypically related diseases such as CRD or LCA; nevertheless, important novel RP-associated genes have not been reported in Japanese since 2017. It is likely that adopting whole exome sequencing or whole genome sequencing would lead to an increased diagnostic rate, particularly in non-RP cases. Lastly, the number of non-RP cases analyzed was too small to reach any conclusions. Nevertheless, the data are sufficient to determine that there is a significant overlap between genes in RP and CRD and that observing this is rare in some genes but more frequent in other genes, such as *CRX*, *PRPH2*, or *RP1*.

In conclusion, our study stringently applied IRD-specific ACMG/AMP rules intended to unify the genetic diagnosis of the disease within Japan in a large number of patients with RP and allied diseases. This allowed us to substantially update the genetic background of Japanese RP patients by detecting many new disease-associated variants. The results may serve as an important foundation for developing future gene- or variant-specific treatments.

## Data Availability

All data relevant to the study are included in the article or uploaded as supplementary information.

## Declarations

### Ethics approval

This study was approved by the ethics committees of all the collaborating hospitals and was conducted in accordance with the tenets of the Declaration of Helsinki on biomedical research involving human subjects. Written informed consent was obtained from all subjects prior to participation in the study.

### Patient consent for publication

Not required.

### Authors’ contributions

KG, YK, MA, YI, K-HS, and KMN designed the study. KG, YK, MA, YMu, MF, KFujiw, KH, MI, THi, KM, MT, JO, AFS, TK, HU, HK, SU, THa, YH, AM, KK, SK, YW, TA, TN, YI, K-HS, and KMN collected the samples and clinical data. KG, YK, HI, MY, ME, KFujit, CT, and YMo performed the experiments and analyzed the data. KG, YK, MA, YMo, K- HS, and KMN contributed to the manuscript preparation and editing. All authors contributed to study conception and design, data interpretation, and approved the final manuscript.

## Acknowledgements

The study was supported by the Japan Retinitis Pigmentosa Registry Project and grants from Japan Agency for Medical Research and Development (23ek0109632 to YI, 23ym0126071h0002 and 23ek0109660h0001 to KMN), Japan Society for the Promotion of Science (20K23005 and 22K16969 to YK, 22K09831 to SU, 21K09756 to THa, 22K09825 to KK, and 23H03059 to KMN), Japanese Retinitis Pigmentosa Society (JRPS) Research Grant to YK, and Takayanagi Retina Research Award to YK. We acknowledge the Laboratory for Genotyping Development in RIKEN and the RIKEN-IMS Genome Platform.

