## Supplementary-Figure for "Disease-specific variant interpretation highlighted the genetic findings in 2325 Japanese patients with retinitis pigmentosa and allied diseases"

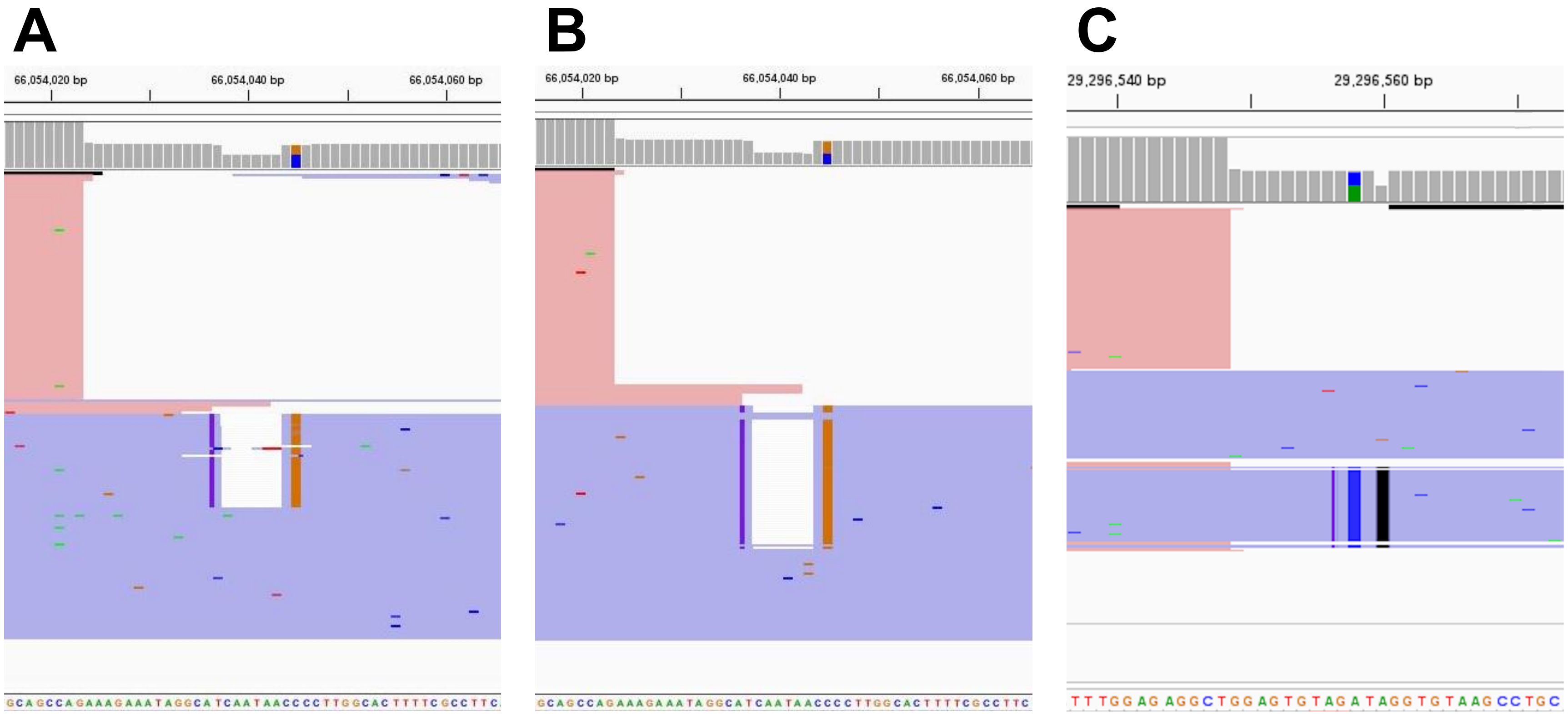


**Figure S1. Integrative Genomics Viewer (IGV) results of variants that corrected the annotations manually.**

We manually corrected the annotations of three variants including two previously reported. The IGV results for [c.1485_1493delinsCGAAAAG in *EYS*] detected in (A) JU-540 and (B) N-303 and [c.568_572delinsAGCAGGCTTACACCAGCAGGCTTT in *PCARE*] detected in (A) N-585-0 are shown. The detailed corrections of three variants are shown in Table S3.
